# Personalization improves the biomechanical efficacy of foot progression angle modifications in individuals with medial knee osteoarthritis

**DOI:** 10.1101/2020.12.15.20248220

**Authors:** Scott D Uhlrich, Julie A Kolesar, Łukasz Kidziński, Melissa A Boswell, Amy Silder, Garry E Gold, Scott L Delp, Gary S Beaupre

**Author notes:** Please direct correspondence to Scott D Uhlrich.

## Abstract

**Objectives:** The goal of this study was to evaluate the importance of personalization when selecting foot progression angle modifications that aim to reduce the peak knee adduction moment in individuals with medial knee osteoarthritis.

**Design:** One hundred seven individuals with medial knee osteoarthritis walked on an instrumented treadmill with biofeedback instructing them to toe-in and toe-out by 5° and 10° relative to their self-selected foot progression angle. We selected individuals’ personalized foot progression angle as the modification that maximally reduced their larger knee adduction moment peak. Additionally, we used lasso regression to identify which secondary changes in kinematics made a 10° toe-in gait modification more effective at reducing the first knee adduction moment peak.

**Results:** Sixty-six percent of individuals reduced their larger knee adduction moment peak by at least 5% with a personalized foot progression angle modification, which is more than (p<0.001) the 54% and 23% of individuals who reduced it with a uniformly-assigned 10° toe-in or toe-out modification, respectively. When toeing-in, greater reductions in the first knee adduction moment peak were related to an increased frontal-plane tibia angle (knee more medial than ankle), a more valgus knee abduction angle, reduced contralateral pelvic drop, and a more medialized center of pressure in the foot reference frame.

**Conclusions:** Personalization increases the proportion of individuals with medial knee osteoarthritis who may benefit from modification of their foot progression angle.

## Introduction

Knee osteoarthritis is a leading cause of disability worldwide^1^ and is accelerated by excessive compressive loading in the joint^2^. The medial compartment of the knee is affected 3.5 times more frequently than the lateral compartment^3^, likely because the majority of compressive knee contact force is transmitted through the medial compartment^4^. Since medial compartment force cannot be directly measured in a native knee, the knee adduction moment (KAM) is commonly used as a surrogate measure of medial loading. The KAM is related to the medio-lateral distribution of loading in the knee^5^ and is also associated with the presence^6^, severity^7^, and progression^8^ of medial compartment knee osteoarthritis. The early-stance KAM peak (i.e., the first peak) is larger than the late-stance KAM peak (i.e., the second peak) in ensemble-averaged curves^9^, but some individuals walk with a larger late-stance KAM peak^10,11^. While the proportion of healthy individuals who walk with a larger second KAM peak has been estimated to be between 15% and 35%, the proportion of individuals with medial knee osteoarthritis who walk with a larger first versus second KAM peak is unknown^10,11^.

Conservative interventions, like modifying the foot progression angle (FPA), can reduce the KAM, but these interventions often prescribe the same modification to all individuals. On average, toe-in reduces the first KAM peak^12^ and toe-out reduces the second KAM peak^13,14^. Six-week gait retraining programs that uniformly assigned individuals with medial knee osteoarthritis to toe-in^15^ or toe-out^16^ have improved knee pain and reduced the first and second KAM peaks, respectively. However, not all participants in these studies reduced the targeted KAM peak, and the effects of the modification on each individual’s larger KAM peak were not reported. Personalization may increase the achievable reduction in KAM for individual subjects^17–20^. For example, healthy subjects who were assigned a personalized FPA modification reduced their larger KAM peak by 19%, which was greater than the 8-11% reductions achieved when all subjects uniformly toed-in or toed-out^10^. The effect of personalization on the ability of FPA modifications to reduce the larger KAM peak in individuals with medial knee osteoarthritis is unclear.

Although the first KAM peak is the larger peak for most individuals, and toeing-in reduces this peak on average, one study found that 41% of healthy individuals with a larger first KAM peak did not reduce it by toeing-in^10^. The varied biomechanical efficacy of toe-in gait may be a result of the varied kinematic strategies that individuals adopt when changing their FPA. When instructed to toe-in, many individuals also change their hip internal rotation angle^20^, ankle inversion angle^21^, tibia angle^22^, and step width^17^. Understanding which secondary kinematic changes are related to greater reductions in the first KAM peak could improve the efficacy of toe-in gait.

Our study examines the importance of personalizing FPA modifications in patients with medial knee osteoarthritis. We first investigated what proportion of individuals with medial knee osteoarthritis walk with a larger first versus second KAM peak. We next evaluated what fraction of this population reduced their larger KAM peak by at least 5% with a personalized FPA modification and compared this to the fraction that reduced it with a uniformly-assigned toe-in or toe-out modification. Finally, we identified which kinematic strategies for adopting a toe-in gait modification were associated with greater reductions in the first KAM peak.

## Methods

### Participants

One hundred seven individuals with medial knee osteoarthritis participated in this study after providing informed consent in accordance with Stanford University Institutional Review Board protocols (Table 1). Individuals were included if they 1) had a medial compartment knee osteoarthritis grade between one and three on the Kellgren-Lawrence scale as determined by a radiologist (GEG) from anterior-posterior weightbearing radiographs, 2) had medial knee pain of three or greater on an 11-point numeric rating scale, 3) were able to walk safely on a treadmill without an ambulatory aid for 25 minutes, and 4) had a BMI less than 35.

**Table 1:** Participant characteristics.

| Characteristics | Mean (SD) |
| --- | --- |
| N | 107 |
| Age (yr) | 59.2 (11.0) |
| Gender | F:64, M:43 |
| Height (m) | 1.70 (0.11) |
| Mass (kg) | 78.6 (14.6) |
| BMI | 27.2 (4.4) |
| Walking Speed (m/s) | 1.20 (0.13) |
| Kellgren & Lawrence Grade | I:27, II:50, III:30 |

### Data Collection

Participants attended two separate visits to a motion capture laboratory. During both visits, nineteen retroreflective markers were placed bilaterally on the 2^nd^ and 5^th^ metatarsal heads, calcanei, medial and lateral malleoli, medial and lateral femoral epicondyles, anterior and posterior superior iliac spines, and the C7 vertebrae. Eighteen additional markers were used to aid in limb tracking. Marker data were collected with an 11-camera optical motion capture system at 100 Hz (Motion Analysis Corporation, Santa Rosa, CA, USA). Ground reaction forces from a split-belt treadmill (Bertec Corporation, Columbus, OH, USA) were synchronously collected at 2000 Hz. Force and marker data were streamed into MATLAB R2015b (The MathWorks, Inc., Natick, MA, USA) for real-time estimation of the FPA^10^. At the beginning of each visit, participants performed a static calibration trial, which we used to estimate knee and ankle joint center locations, joint axes of rotation, and tracking marker reference locations^10^. Participants were then instructed to circumduct each leg for the estimation of hip joint center locations from thigh and pelvis kinematics^23^.

During the first lab visit, participants acclimated to walking on the treadmill and received feedback to modify their FPA. They first walked naturally for 10 minutes at a self-selected speed, then performed two 10-minute FPA modification trials where they received real-time vibrotactile feedback instructing them to toe-in and toe-out by 10° relative to their baseline FPA. Two vibrotactile motors (Model: C2; Engineering Acoustics, Inc., Casselberry, FL, USA) affixed to the medial and lateral aspects of the proximal tibia provided feedback at the end of stance phase following any step where the FPA was not within 2.5° of the trial’s target angle^10^. Feedback was only given to the more symptomatic leg, but participants were encouraged to match the FPA modification with their contralateral limb.

During the second lab visit, participants warmed up on the treadmill for five minutes, then performed a two-minute baseline walking trial. Participants next received feedback while they practiced walking with 5° and 10° of toe-in and toe-out for a minimum of one minute at each angle or until the participant reported being able to walk comfortably with the modification. FPA evaluation trials were then performed in random order for each of the four modifications for a minimum of two minutes or until the FPA of 30 steps fell within the target ±2.5° range.

### Data Analysis

Motion and force data were low-pass filtered at 15 Hz using a zero-lag, 4^th^ order, Butterworth filter in MATLAB. The KAM was defined as the frontal-plane component of the 3-dimensional knee moment, expressed in a proximal tibial reference frame as a percentage of bodyweight and height (%BW* ht). The knee moment was calculated as the cross product of the ground reaction force and the vector from the knee joint center to the center of pressure^10^. The first and second KAM peaks were identified as the maximum value of the KAM timeseries during the first and second halves of the stance phase, respectively. To quantify the relative magnitude of the KAM peaks during the baseline trial, we computed the percent difference between KAM peaks by subtracting the first peak from the second peak, then dividing by the larger of the peaks. We analyzed the final 20 steps from the baseline trial and the final 20 steps from each modified FPA evaluation trial for which the FPA was within 2.5° of the trial’s target angle. The 20 analyzed steps from the modified FPA trials were divided into two sets of 10. The average KAM peak from the first set of 10 steps of each trial was used to select the personalized FPA as the 5° or 10° FPA modification that maximally reduced the larger KAM peak as measured at baseline. The average KAM peak from the second set of 10 steps from each trial was used to compare reductions between personalized and uniformly-assigned FPA modifications. A 5% reduction in the KAM peak was considered clinically meaningful based on previous cohorts who reduced pain with similar reductions in the KAM peak^24,25^.

We used a linear regression model with lasso (L1) regularization to identify the subset of kinematic changes that explained the most variance in first KAM peak reductions from a 10° toe-in modification. Inputs to the model were changes in kinematic features, with established relationships to the KAM, from the baseline trial to the 10° toe-in trial (Table 2). Unless otherwise noted, kinematic timeseries variables were reduced to scalar inputs by selecting the timeseries value at the time of the first KAM peak. Trunk sway^12^, knee flexion and abduction angle^26^, pelvic list and rotation angle^27^, frontal-plane tibia angle^22^, FPA^10^, step width^10^, and distance between knee joint centers^12^ were computed using marker positions. Trunk sway was computed using the midpoint of the posterior superior iliac spine markers and the C7 marker^22^; the scalar value was the maximum angle during stance, with a positive angle representing trunk lean over the stance leg. Knee angles were computed using the positions of the lateral femoral epicondyle and the hip, knee, and ankle joint centers^26^; positive angles represent flexion and abduction (valgus). Pelvic list and axial rotation were computed as the angle between the medio-lateral laboratory axis and the line connecting the anterior superior iliac spines projected into the laboratory frontal and transverse planes, respectively; positive angles represent a more superior and anterior stance side of the pelvis. The frontal-plane tibia angle was computed as the angle between the laboratory vertical axis and the line connecting the knee and ankle joint centers projected into the laboratory frontal plane^22^; a positive angle represents the knee medial to the ankle. Step width was computed as the medio-lateral distance between the right and left midfoot during consecutive steps and the scalar value was the distance at 50% of the stance phase. Using anthropometric measurements of the foot^28^, we defined the midfoot as the point 70% of the distance from the calcaneus to the 2^nd^ metatarsal head marker along the line connecting the markers. The medio-lateral distance between knee joint centers and centers of pressure were computed from consecutive steps^10^. We determined the center of pressure position relative to the foot by first defining an anatomical foot reference frame when the foot was flat on the ground during the static calibration trial. The frame origin was the calcaneus marker and it had the following axes: the line from the calcaneus to the second metatarsal head marker projected onto the laboratory transverse plane, the laboratory vertical axis, and the mutual perpendicular axis. During gait, the center of pressure location in the foot reference frame was defined as the point where the vector originating at the center of pressure and pointing along the laboratory vertical axis intersected the foot transverse plane. Center of pressure positions medial to the anterior-posterior foot axis and anterior to the calcaneus were considered positive.

**Table 2:**
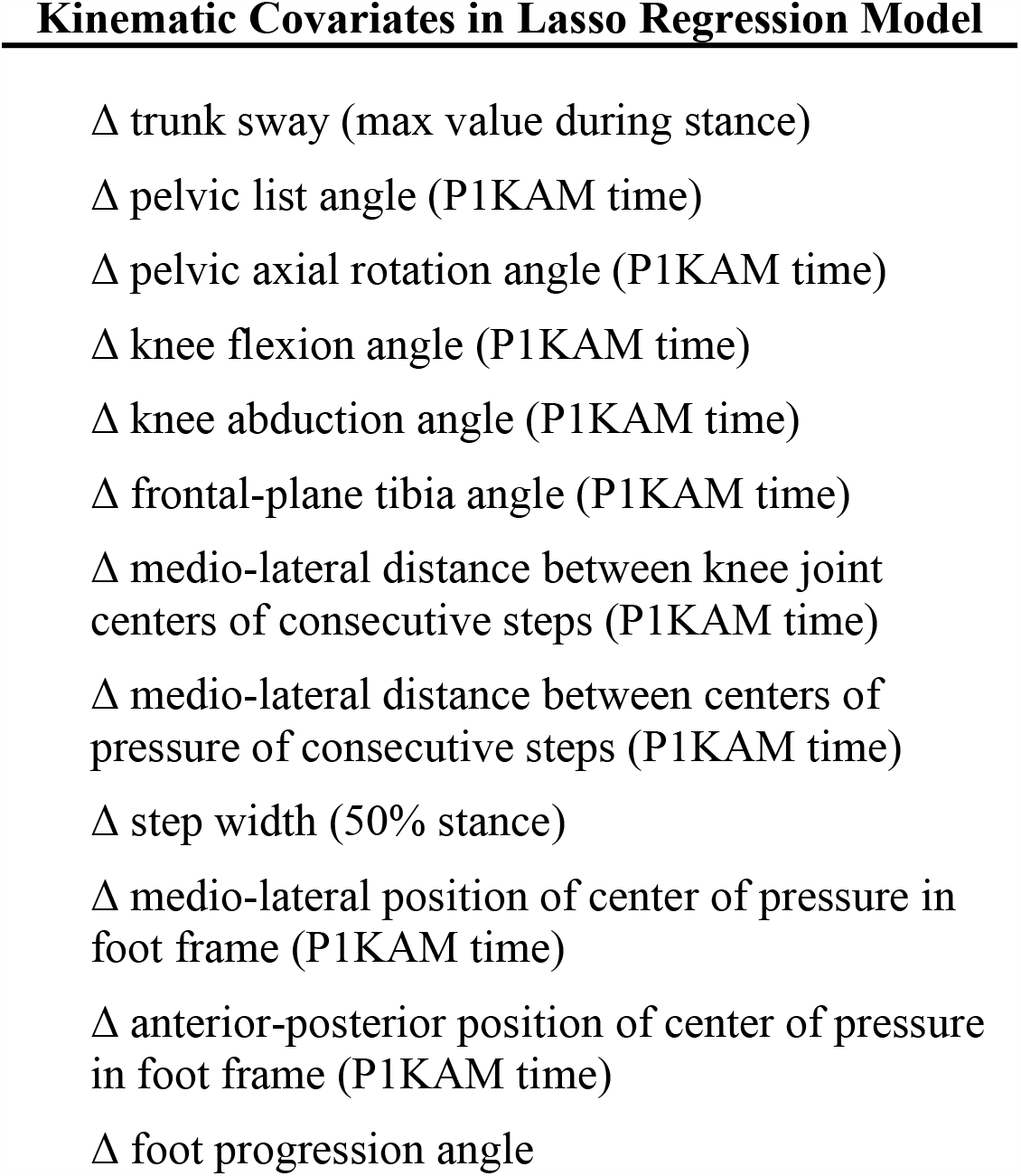
Kinematic features related to the knee adduction moment (KAM) were used as covariates in a linear regression model with lasso regularization to predict changes in the first KAM peak (P1KAM) when participants toed-in by 10°. All covariates were the change (Δ) in scalar value from the baseline to the 10° toe-in trial. The time when the timeseries was evaluated is noted in parentheses.

### Statistics

The proportion of participants who reduced their larger KAM peak by at least 5% with a personalized FPA modification was compared to the proportion who reduced this peak when all participants were assigned a uniform 10° FPA modification using a mid-p-value McNemar’s test of proportions. This test was performed in MATLAB (R2015b). Changes in the larger KAM peak between modifications were compared using a Wilcoxon signed-rank test, and the median and 95% confidence interval (CI) of the differences are reported. These tests were performed in R^29^ (v. 3.5.3, R Foundation for Statistical Computing, Vienna, Austria). Statistical significance was set to 0.05 and p-values are reported with a Bonferroni correction for multiple comparisons.

Lasso regularization was used to select a reduced set of kinematic features that explained the most variance in first KAM peak reductions^30^. The lasso penalty coefficient, λ, was selected after performing 10 iterations of 10-fold cross-validation using the *glmnet* package^31^ in R^29^ (v. 3.5.3). We used the one-standard-error rule for selecting lambda: for each iteration, the λ value that yielded a prediction error that was one standard error greater than the minimum prediction error was selected, thereby removing more features than the λ that minimized prediction error. We used the average lambda value from cross-validation for the final regression model with regularization. Post-selection p-values (α=0.05) and 95% confidence intervals for model coefficients conditional on the lasso selection were computed using the *selectiveInference* package in R^32–34^. We report coefficients (β) and r^2^ (square of Pearson’s r) values from an unregularized linear model with the lasso-selected features as covariates and a reduction in KAM considered positive. For this model, we report coefficients with both standardized (zero mean and unit standard deviation) and unstandardized inputs. To display the differences in the toe-in kinematic change variables that were selected by the lasso, the kinematic timeseries were averaged for the participants above the 90^th^ percentile of first KAM peak reduction (i.e., KAM reducers) and below the 10^th^ percentile (i.e., KAM non-reducers).

## Results

At baseline, 100 of 107 participants (93%) walked with a larger first versus second KAM peak (Figure 1). Seventy-one participants (66%) reduced their larger KAM peak by at least 5% with a personalized FPA modification (Figure 2, Table S1). This was greater than (p<0.001) the number of participants who reduced their larger KAM peak when everyone toed-in by 10° (58 participants, 54%) and toed-out by 10° (25 participants, 23%). Conversely, 11 participants (10%) *increased* their larger KAM peak by more than 5% when everyone toed-in by 10°, and 47 participants (44%) *increased* it when everyone toed-out by 10°. With a uniformly-prescribed FPA modification, some individuals *increased* their larger KAM peak by as much as 35% (Figure 2, Table S1). Of the 71 participants who reduced their larger KAM peak with a personalized FPA, 11 maximally reduced it with 5° toe-in, 42 with 10° toe-in, 7 with 5° toe-out, and 11 with 10° toe-out.

**Figure 1:**
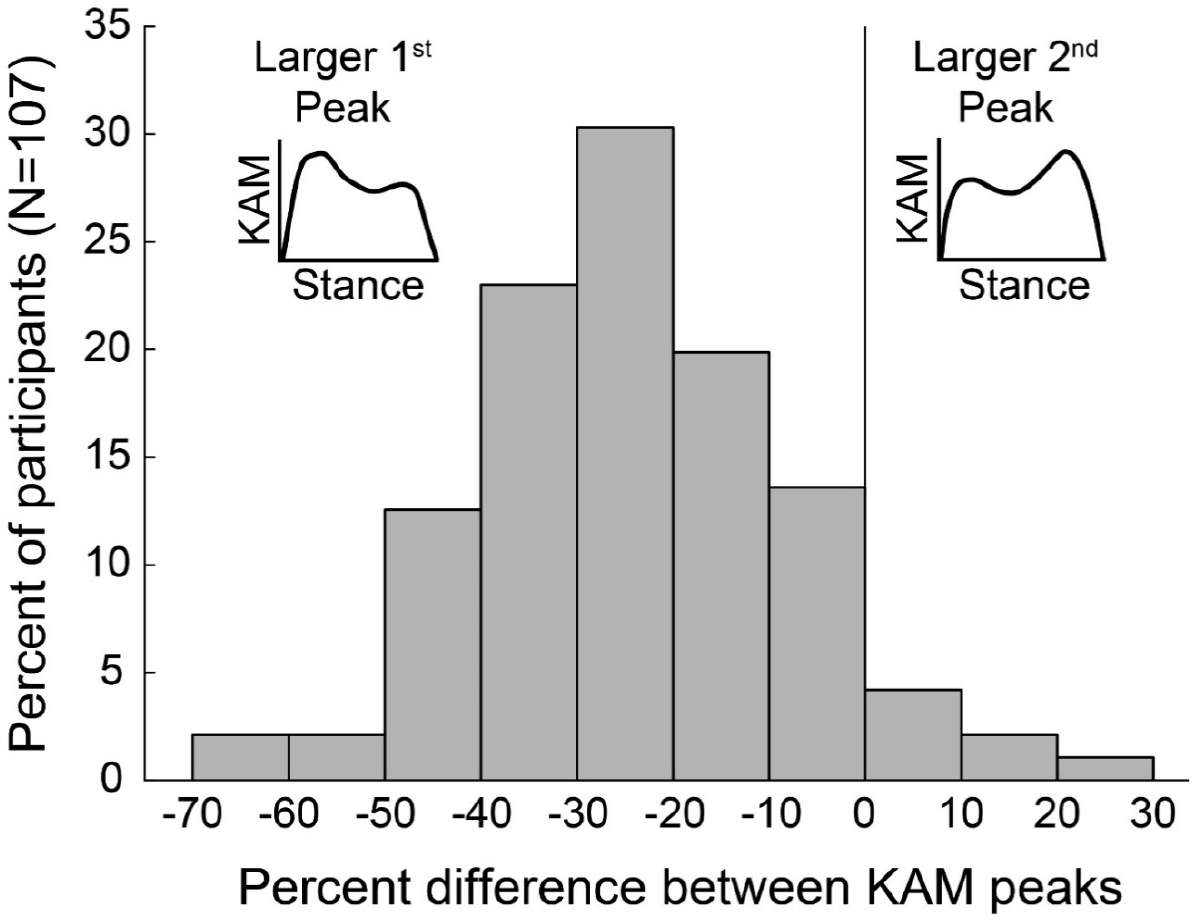
100 of 107 individuals (93%) walked with a larger first knee adduction moment (KAM) peak at baseline, while the remaining seven walked with a larger second KAM peak.

**Figure 2:**
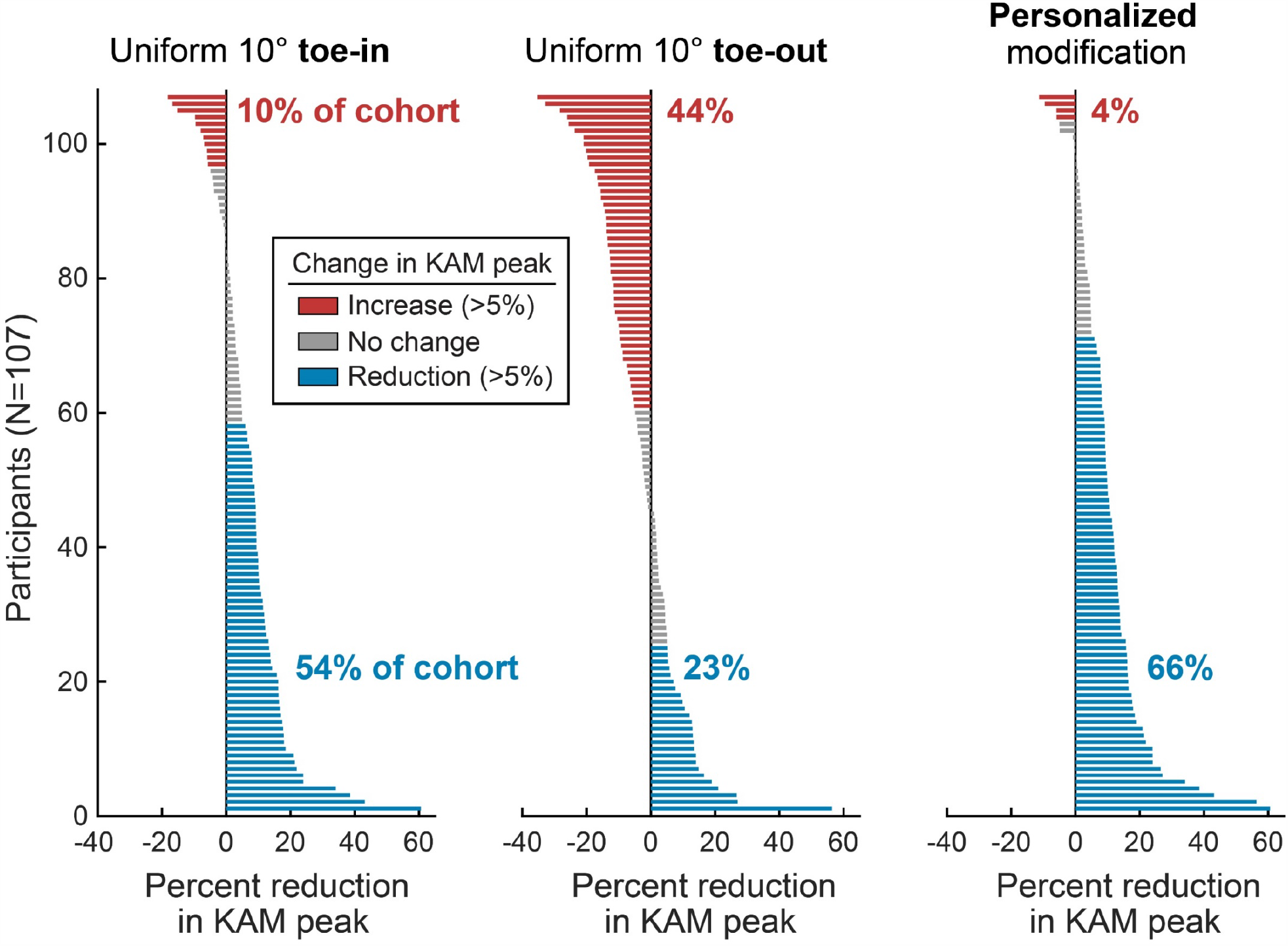
Changes in each individual’s larger knee adduction moment (KAM) peak when all participants toed-in by 10°, toed-out by 10°, and walked with their personalized foot progression angle modification. A greater proportion (p<0.001) of participants reduced (shown as positive) their larger KAM peak by at least 5% (blue) with a personalized modification (66%) compared to a uniformly-assigned toe-in (54%) or toe-out (23%) modification. Importantly, 10-44% of individuals *increased* their larger KAM peak by more than 5% (red) when uniformly assigned a 10° foot progression angle modification, with some individuals increasing their larger KAM peak by as much as 35%.

Among all participants, a personalized FPA modification reduced their larger KAM peak by an average of 11±11% (0.31±0.28 %BW* ht), which was greater than the 7±11% (0.21±0.30 %BW* ht) reduction when all individuals toed-in by 10° (median difference: 0.09 %BW* ht, 95% CI: 0.08-0.10 %BW* ht, p<0.001), and the 3±14% (0.09±0.38 %BW* ht) increase when all individuals toed-out by 10° (median difference: 0.40 %BW* ht, 95% CI: 0.38-0.42 %BW* ht, p<0.001). Additionally, a personalized approach allows for an intervention to only be prescribed to individuals for whom an FPA modification reduces their KAM. For example, among the 66% of individuals who reduced their larger KAM peak by at least 5% with a personalized FPA modification (Figure 2), the average reduction was 16±10% (0.45±0.22 %BW* ht).

The lasso regression model selected four of the original 12 kinematic covariates to explain how effectively a 10° toe-in modification reduced the first KAM peak (Table 3, Figure 3). An unregularized linear model using these four covariates explained the variance in the first KAM peak reduction with adjusted r^2^=0.42. When adopting a toe-in gait modification, greater reductions in the first KAM peak were related to an increased frontal-plane tibia angle (knee more medial than ankle, p<0.001, Figure 3a), an increased knee abduction angle (more valgus, p<0.001, Figure 3b), a smaller pelvic list angle (less contralateral pelvic drop, p=0.027, Figure 3c), and a smaller lateral shift in center of pressure in the foot frame (p=0.004, Figure 3d).

**Table 3:**
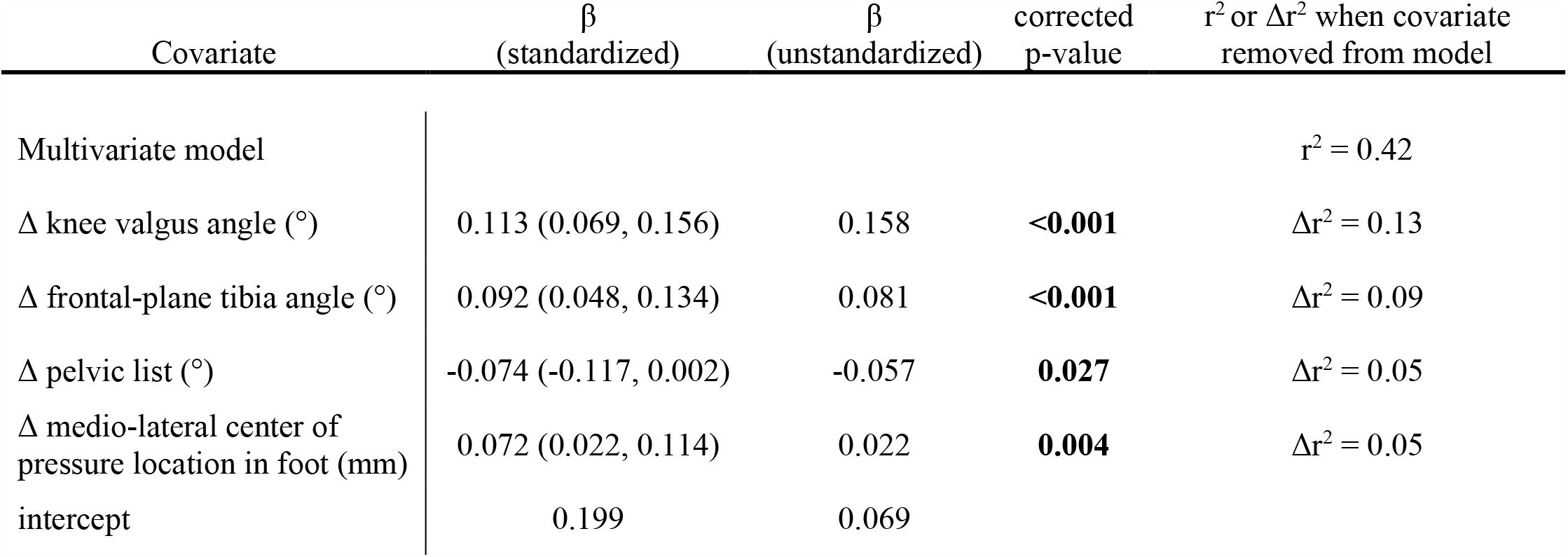
The four covariates selected by the lasso are used in an un-regularized model to explain changes in the first KAM peak when toeing-in by 10° compared to baseline. The Δ represents a change in a kinematic parameter between baseline and toe-in walking, and a reduction in the first KAM peak is considered positive. 95% confidence intervals are presented for the standardized coefficients.

| Covariate | $\beta$<br>(standardized) | $\beta$<br>(unstandardized) | corrected<br>p-value | $r^2$ or $\Delta r^2$ when covariate<br>removed from model |
| --- | --- | --- | --- | --- |
| Multivariate model | | | | $r^2 = 0.42$ |
| $\Delta$ knee valgus angle ($^\circ$) | 0.113 (0.069, 0.156) | 0.158 | **<0.001** | $\Delta r^2 = 0.13$ |
| $\Delta$ frontal-plane tibia angle ($^\circ$) | 0.092 (0.048, 0.134) | 0.081 | **<0.001** | $\Delta r^2 = 0.09$ |
| $\Delta$ pelvic list ($^\circ$) | -0.074 (-0.117, 0.002) | -0.057 | **0.027** | $\Delta r^2 = 0.05$ |
| $\Delta$ medio-lateral center of<br>pressure location in foot (mm) | 0.072 (0.022, 0.114) | 0.022 | **0.004** | $\Delta r^2 = 0.05$ |
| intercept | 0.199 | 0.069 |  |  |

**Figure 3:**
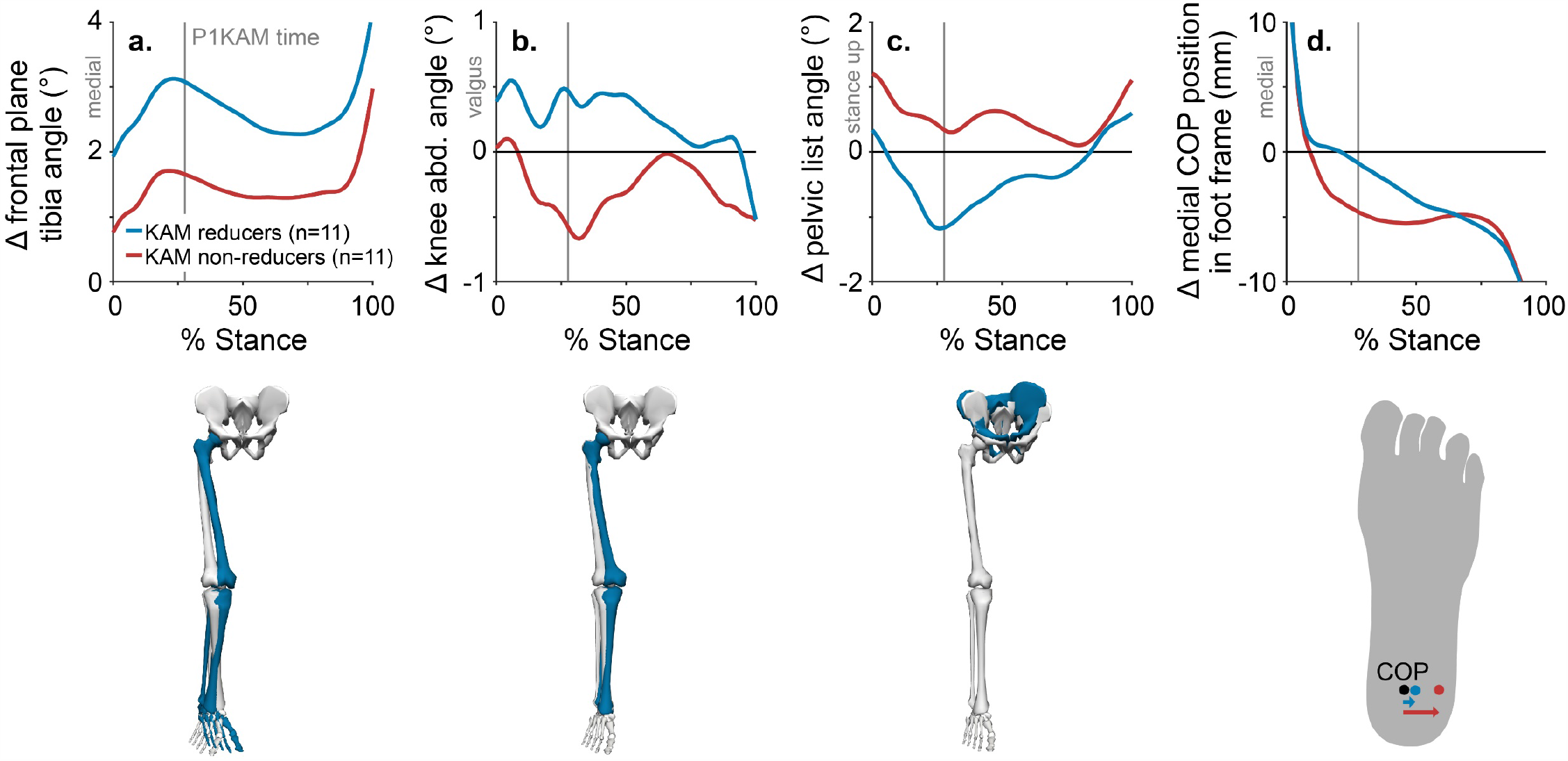
Compared to baseline walking, toeing-in with a more medialized frontal-plane tibia angle (**a**), a more valgus knee abduction (abd.) angle (**b**), an elevated swing-side pelvis (i.e., more negative pelvic list angle), **c**), and a more medialized center of pressure (COP) in the foot frame (**d**) were associated with greater reductions in the first knee adduction moment peak (Table 3). Changes in kinematic timeseries from baseline to 10° toe-in are shown for individuals above the 90^th^ percentile of KAM reduction (KAM reducers, blue) and for individuals below the 10^th^ percentile of KAM reduction (KAM non-reducers, red). To generate scalar inputs for the linear model (Table 3), we selected the timeseries values at the time of the first KAM peak (P1KAM).

## Discussion

This study demonstrates the importance of personalizing foot progression angle modifications in order to maximize the number of individuals with medial knee osteoarthritis who can reduce their knee adduction moment peak with a modification. Ninety-three percent of our participants walked with a larger first knee adduction moment peak. Although a toe-in gait modification reduces the first knee adduction moment peak on average^12^, only 54% of the participants in our study reduced their larger knee adduction moment peak when toeing-in by 10°. This was notably less than the 66% who reduced it with a personalized toe-in or toe-out modification.

Personalization not only enables a greater proportion of individuals to reduce their KAM peak, but perhaps more importantly, it also avoids *increasing* the KAM peak in a portion of the population. Previous studies that uniformly assigned KAM-reducing interventions to all participants reported that 18-33% of participants did not reduce the targeted KAM peak^16,25^, with some participants increasing it by as much as 10%. Ten to forty-four percent of our participants increased their KAM peak by more than 5% with a uniformly-assigned 10° FPA modification, with some individuals increasing their KAM peak by as much as 35%. Greater reductions in the KAM from an intervention are correlated with greater improvements in pain and function^24^, suggesting that the clinical efficacy of load-reducing interventions may be improved by only prescribing them to the subset of patients for whom the intervention reduces their KAM. Selecting patients who are most likely to benefit and selecting the intervention that maximizes their KAM reduction may reduce variance in clinical trial outcomes and improve our understanding of how these tools fit into the conservative management of knee osteoarthritis.

Two recent randomized controlled trials that investigated personalized and non-personalized gait retraining techniques reported different levels of clinical efficacy^35,36^. Cheung et al. gave patients visual biofeedback of the KAM curve and verbally suggested several kinematic modifications to help reduce their KAM peak^36^. Hunt et al. uniformly assigned toe-out to all participants^35^. At follow up, participants in the KAM biofeedback study reduced their KAM peak by 25% and improved their WOMAC pain by 59%, while participants in the toe-out study reduced their second KAM peak by 7% and improved their WOMAC pain by 33%. The KAM biofeedback study represents a highly personalized approach. Providing participants with direct, continuous feedback of the KAM likely enhanced the KAM and pain reductions; however, clinical translation of real-time biofeedback of the KAM curve is challenging since it requires a gait analysis laboratory. The toe-out study was not personalized, potentially limiting the degree of KAM reduction that each participant achieved; however, its training technique of instructing participants to match their FPA to a piece of tape on a mirror is more easily translated into the clinic or home. Our proposed personalization protocol could leverage the strengths of both studies. A single visit to a gait laboratory could identify which, if any, FPA modification maximally reduces an individual’s larger KAM peak, and subsequent gait retraining visits could be performed at home or in the clinic^35,37^.

Forty-four percent of our cohort who walked with a larger first KAM peak did not reduce it by at least 5% when toeing-in by 10°; however, our lasso regression model identified kinematic strategies for adopting toe-in gait that could make it more effective in future studies. Instructing patients to toe-in with a medialized tibia angle, a more valgus knee angle, a more medialized center of pressure, and less contralateral pelvic drop may help them reduce their first KAM peak. All of these kinematic strategies have previously been suggested as single-parameter gait modifications for reducing the KAM^22,27,38,39^. Giving biofeedback on all four of these kinematic changes along with the FPA may result in larger KAM peak reductions than giving biofeedback on the FPA alone^17,40,41^, but modifying a single parameter is preferable for patient learning and compliance^15^. To retain the simplicity of a single-parameter gait modification, these kinematic strategies for toeing-in could be verbally suggested alongside FPA biofeedback during gait retraining sessions to improve a patient’s ability to reduce their first KAM peak.

The lasso regression results also provide insight into the mechanisms by which toe-in reduces the first KAM peak. Toe-in has been suggested to reduce the KAM lever arm during early stance by either increasing the tibia angle through hip internal rotation and knee flexion, lateralizing the center of pressure relative to the pelvis, or both^12^. In our study, the change in tibia angle explained the most variance of any covariate in the model, while the medio-lateral distance between the centers of pressure from consecutive steps was not selected by the lasso. Interestingly, the two variables that explained the most variance in the model, the change in tibia angle and the change in knee abduction angle, both relate to frontal-plane limb alignment. Although they are similar, the tibia angle is expressed in the laboratory frame, so it can be changed with a variety of multi-joint kinematic changes. The knee abduction angle, however, is expressed in the knee reference frame, making it specific to changes in the frontal-plane knee angle. Since both variables explained unique variance in the model, individuals likely altered their KAM lever arm with one or both of these strategies. Finally, all four covariates selected by the lasso relate to medial thrust gait, which involves medializing the knee and elevating the swing-side pelvis^27^ and has been observed to cause a medial shift in the center of pressure in the foot reference frame^42^. Together, these results suggest that when a toe-in gait modification is most effective, it may be reducing the first KAM peak with a similar mechanism as medial thrust gait.

Our study has several limitations. First, we demonstrated the importance of personalizing FPA modifications for maximizing the number of individuals who can reduce their larger KAM peak, but we did not study the effects of personalization on clinical outcomes such as pain and function. Studies to assess these important clinical outcomes are underway. Second, the need to personalize gait modifications complicates clinical translation. Traditionally, measuring the KAM requires a force plate, motion capture, and a trained gait analyst. The KAM has been computed outside of the gait laboratory, however, using inertial measurement units with and without force-instrumented shoes^43,44^, as well as with machine learning models that use ground reaction forces or simplified kinematic measures as inputs^45–47^. Further work is necessary to validate the ability of these technologies to accurately select personalized modifications. Finally, there is a strong body of literature linking the KAM peak to medial knee osteoarthritis clinical outcomes, but it may not be the optimal mechanical target for disease-modifying interventions. The area under the KAM curve (i.e., the KAM impulse) provides information about cumulative loading, and some studies report that it is a better predictor of clinical outcomes^24,48^ than the KAM peak. Additionally, the KAM does not capture changes in muscle force that occur from the secondary kinematic and coordination changes^10,21,49,50^ that often accompany FPA modifications. Further work comparing the relationships between medial knee osteoarthritis progression and the KAM peak, KAM impulse, or simulation-estimated medial contact force would help clarify the optimal target for conservative interventions that alter joint mechanics.

This study highlights the importance of a personalized approach for prescribing gait modifications to individuals with knee osteoarthritis. Gait modifications may become an important tool for improving symptoms and slowing disease progression, but we and others have shown that not all individuals benefit from a uniformly-prescribed modification. Our study shows that personalization can improve the biomechanical efficacy of gait modifications, suggesting that load-modifying interventions for osteoarthritis should be personalized in future clinical trials and clinical practice. This approach both selects the patients who are most likely to benefit from the intervention and selects the intervention that maximally offloads each individual’s joint. By leveraging rapid improvements in mobile sensing technology, clinicians may soon be able to prescribe personalized gait modifications in a clinical setting, potentially making them an effective tool for the treatment of knee osteoarthritis.

## Data Availability

The dataset and lasso regression code are available at: https://github.com/suhlrich/ToeInKAMReduction.

## Acknowledgements

We would like to acknowledge Healey Montague-Alamin, Brittany Presten, Madeleine Berkson, Naomi Edouard, and Dominic Willoughby for assistance with data collection and processing. We would also like to thank Trevor Hastie and Jonathan Taylor for their assistance with statistical analysis. SDU and MAB were funded by fellowships from the National Science Foundation (DGE-114747, DGE-1656518) and the Stanford Office of the Vice Provost for Graduate Education. This work was also supported by grant P41 EB027060 from the National Institutes of Health and by Merit Review award I01 RX001811 from the United States Department of Veterans Affairs Rehabilitation Research and Development Service. The contents of this work do not represent the views of the Department of Veterans Affairs or the United States Government.

## Author contributions

SDU, JAK, AS, GEG, SLD, and GSB conceived of the study. SDU, JAK, and AS collected the experimental data. GEG graded the radiographs. SDU and MAB analyzed the data, and LK provided statistical expertise. SLD and GSB obtained funding for the study. SDU and JAK drafted the manuscript, and all authors critically revised it for intellectual content. All authors approved the final manuscript before submission. SDU takes responsibility for the integrity of this work.

