## Supplemental Materials for "Personalization improves the biomechanical efficacy of foot progression angle modifications in individuals with medial knee osteoarthritis"

**Supplemental Material**

**Table S1**: The number of participants who increased or reduced their larger knee adduction moment peak (P1KAM or P2KAM) by at least 5% with uniform and personalized FPA modifications. Participants who did not change their knee adduction moment peak by at least 5% are not reported. Reported p-values compare the proportion who reduced their PKAM with uniform modifications to a personalized modification.

| | Baseline | Reduced PKAM with 10° toe-in | Increased PKAM with 10° toe-in | Reduced PKAM with 10° toe-out | Increased PKAM with 10° toe-out | Reduced PKAM with personalized FPA |
|---|---|---|---|---|---|---|
| Larger P1KAM | 100 | 56 (56%) | 10 (10%) | 20 (20%) | 47 (47%) | 66 (66%) |
| Larger P2KAM | 7 | 2 (29%) | 1 (14%) | 5 (71%) | 0 (0%) | 5 (71%) |
| Total | 107 | 58 (54%, p<0.001) | 11 (10%) | 25 (23%, p<0.001) | 47 (44%) | 71 (66%) |
